# A Qualitative Evidence Synthesis Exploring Caregiver Information Needs and Experiences Caring for a Child with Chronic Heart Failure

**DOI:** 10.1101/2024.02.14.24302803

**Authors:** Chentel Cunningham, Katie Schroeder, Tabatha Plesuk, Jennifer Conway, Mark Haykowsky, Shannon D. Scott

**Author notes:** **Corresponding Author:** Shannon D. Scott, 5-187 Edmonton Clinic Health Academy, 11405 – 87 Ave NW, Edmonton, AB Canada, T6G 1C9. Funding Sources: Canadian Institute of Health Research (CIHR) Canadian Nurses Foundation Alberta Registered Nurses Educational Trust (ARNET) Women’s and Children’s Health Research Institute (WCHRI) University of Alberta, Faculty of Nursing University of Alberta, Faculty of Graduate Studies and Research. Ethical Considerations: None.

## Abstract

**Aim:** To synthesize qualitative studies relating to caregivers’ information needs and experiences caring for a child with chronic heart failure.

**Background:** Children with chronic heart failure (HF) place a considerable burden on healthcare systems each year and is associated with significant stress for caregivers. Despite substantial knowledge generation and implementation amongst healthcare providers in this field, knowledge translation strategies that target caregiver audiences have lagged. To date, little is known about what literature exists about caregiver’s information needs and experiences caring for a child with HF.

**Design:** Qualitative evidence synthesis

**Methods:** Sandelowski and Barroso’s Handbook for Synthesizing Qualitative Research guided our review, cross-referencing the ENTREQ statement to report our findings. With librarian guidance, seven databases were searched in November 2023. Search results were imported into Microsoft Excel from EndNote 21. Two independent reviewers (CC, KS), supervised by a senior author (SDS), conducted the initial title/abstract and secondary full-text screens.

**Results:** One study met the inclusion criteria outlined in our synthesis, identifying three themes and 10 subthemes. Our findings impeded the ability to conduct any analysis.

**Conclusion:** This review was the first of its kind, looking to synthesize caregiver’s information needs and experiences caring for a child with chronic HF. One article met the inclusion criteria. This synthesis highlights a significant knowledge gap in caregivers’ experiences and information needs when caring for a child affected by chronic heart failure. This study emphasizes that previous educational tools were not designed using research evidence about caregiver experience. This also validates the critical need for qualitative interviews with caregivers about their information needs and experiences to provide more relevant and understandable information to caregivers in this difficult situation.

**Reporting Method:** EQUATOR guidelines, following the ENTREQ statement.

**Patient Contribution:** Caregivers of children with heart failure were the population in this qualitative evidence synthesis.

**What does this paper contribute to the wider global clinical community?:**

- To our knowledge, this is the first qualitative evidence synthesis about caregiver information needs and experiences caring for a child with HF.
- One included study was identified, highlighting a significant knowledge gap and justifying the need for the next step in our research, employing qualitative interviews in this context.
- Healthcare professionals who care for this vulnerable patient population have very limited literature to base practice.
- Further research about caregivers’ information needs and experience in pediatric chronic heart failure context is needed to improve care provision to these vulnerable children,

## INTRODUCTION

Heart failure (HF) in children is documented to be a complex and burdensome disease.^1-5^ Hospitalizations in the United States are estimated to occur in 11,000-14000 children annually.^1^ Adult HF is better characterized as limited data on incidence and prevalence exists in childhood HF due to small numbers, varying phenotypes, and lack of an overall standardized definition.^2^ It is broadly defined as a clinical and pathophysiological syndrome resulting from ventricular dysfunction, volume or pressure overload, in combination or alone.^7^ The cardinal symptoms of children with HF, no matter the etiology, are poor growth, dyspnea, and anasarca.^3-5^ Diverse etiologies of pediatric HF require specific treatments, each with their own distinct trajectories.

One subset of children with heart failure experience heart muscle disease that results in systolic and diastolic dysfunction, resulting in a more chronic HF phenotype.^3^ This makes for an uncertain, life-long trajectory, with constant burdensome symptom management for families. Examples of conditions that fall under this spectrum include cardiomyopathies, myocarditis, toxin-induced heart failure, genetic/metabolic diseases, and nutritional and neuromuscular conditions. This cohort of children typically only have a limited number of surgical options to relieve them of their HF symptoms (e.g., ventricular assist devices and cardiac transplantation)^2,6^ which are sometimes not considered due to the underlying progressive systematic disease. The goals of care are to manage and control symptoms through complex medical therapies.^5^ Furthermore, these etiologies encompass a population of children who have HF symptoms that present later in childhood from being previously healthy, which lends to a unique family experience compared to most children born with congenital heart disease (CHD) who are diagnosed in-utero or shortly after birth.

A second HF phenotype includes children with congenital heart disease. HF symptoms result from structural abnormalities^2^ and generally present with a more acute presentation from pressure overload or over-circulation.^4^ Treatment strategies for this cohort of children are typically surgical approaches (e.g., correction vs. palliation).^6^ Children with CHD also experience different clinical trajectories compared to children with chronic HF due to over-circulation, which is typically relieved with surgical correction.^7^ However, a small number of children within the CHD population do experience chronic HF due to systolic or diastolic dysfunction, which is categorized under the spectrum of chronic HF.^8^

Improved outcomes for children with chronic HF are occurring due to earlier recognition and advancements in evidenced-based treatment strategies that are the result of increased collaboration among healthcare professionals who specialize in children’s heart failure.^3,4^ These initiatives have led to more children with HF surviving and being discharged into the outpatient setting. Despite these encouraging clinical gains, knowledge translation strategies have not kept pace for the caregiver audiences, bringing about knowledge gaps among caregivers responsible for their child’s complex care.^7^ Within the congenital heart disease realm, there is a distinct call to improve knowledge translation strategies that target caregivers,^10,11^ which has not been documented in the chronic HF population.

When limited information about a child’s disease or treatments is not translated into understandable formats for caregivers, feelings of stress, anxiety and issues with adherence often result.^8,9^ However, when caregivers or caregivers have access to understandable, evidenced-based information, improvements in their confidence and decision-making occur while also reducing healthcare costs.^7^ Access to information tailored to caregivers’ knowledge, needs and experiences is key to developing these effective educational tools, promoting improved adherence to complex treatment strategies and fostering resilience among caregivers to withstand a long, complex medical journey better.^10^ These benefits are congruent for caregivers who have a child affected by chronic HF.

## 1.0 AIM

The aim was to describe and synthesize all available qualitative knowledge related to caregivers’ information needs and experiences caring for a child with chronic HF.

## 2.0 METHODS

### 2.1 Design

Sandelowski and Barroso’s (2007) Handbook for Synthesizing Qualitative Research guided our synthesis and reported according to the ENTREQ statement.^11,12^ With a surge of qualitative research over the last 20 years,^11^ adhering to both these guidelines provided a structured yet flexible means to robustly synthesize an increasing area of knowledge in the field of pediatric cardiology care that can inform and shape nursing practice in an area that is gaining traction in the clinical realm. This study is unique because no qualitative evidence syntheses with the same focus are registered in the Cochrane Library or PROSPERO database.

### 2.2 Search Strategy

Identifying key concepts, search strategy development, and inclusion/exclusion criteria for our qualitative synthesis was informed by the structure outlined in the population, intervention, comparison, outcomes and study type (PICOS) tool.^13^ As Methley et al. (2014) suggested, the PICO tool does not accommodate terms relating to qualitative research. The PICOS tool was used as an alternative to capture the qualitative study design to ensure a comprehensive search, as it is more sensitive than other tools such as SPIDER.

#### Population

Studies were included if they had caregivers who cared for a child with a diagnosis of HF in the home or hospital setting. Chronic HF is defined as a clinical and pathophysiologic syndrome resulting from ventricular dysfunction (systolic or diastolic), with characteristic signs and symptoms (poor growth, feeding difficulties, respiratory distress, exercise intolerance, and fatigue, and is associated with circulatory, neurohormonal, and molecular abnormalities)^2^ that have very limited surgical option for correction (e.g., cardiac mechanical assist device (VAD) or transplant). A caregiver is defined as a biological caregiver, relative or guardian over 18 years of age who is primarily responsible for the daily management of a child between the ages of 0-21 diagnosed with chronic HF by a pediatric cardiologist.

#### Intervention

Intervention relates to the act of providing care to a child diagnosed with chronic HF from an acquired, congenital heart defect or cardiomyopathy etiology. Studies that focused on caregivers caring for a child with congenital heart disease were automatically carried into the second full-text screening phase to screen the article for references to chronic heart failure symptoms. Studies were excluded with CHD if the child’s lesion was amendable to surgical correction or if heart failure was not explicitly stated as part of their clinical course. The care provided could be documented to take place in either the home setting or hospital setting. The intervention includes physical, psychological, emotional, financial, or medical support. Any studies related to children on mechanical heart support (e.g., VADs) were also excluded as they are often a means to controlling advanced heart failure symptoms and have other complex clinical considerations.

#### Comparison

The comparison group for this research question is identified as caregivers’ experiences and knowledge needs in caring for a healthy child compared to a child with HF. Alternatively, there would be no comparison for caregivers with no other children.

#### Outcomes

Outcomes were classified as studies reporting caregivers’ information needs and experiences relating to caring for a child with chronic HF. Caregiver experiences included their participation in care, perception, health information-seeking behaviours, and attitude towards their child’s HF.

#### Study Type

The study type refers to all qualitative methodological approaches (e.g., phenomenological, ethnographic, narrative, grounded theory, etc.).^13^ All studies were included in the title and abstract screen if they were of a qualitative methodology.

### 2.3 Information Sources and Comprehensive Search

The search was conducted in November. 2023 in seven medical, psychological, and socially based databases (Ovid, combining searches from MEDLINE, Scopus, EMBASE, PsycINFO; and a second combined search in Cumulative Index to Nursing and Allied Health Literature (CINHAL), Educational Resources Information Centre (ERIC), & Education Research Complete). All references were imported into a software reference Manager (EndNote 21). To ensure a comprehensive and exhaustive search, consultation with two individuals with an extensive library science background occurred before the search (MK, John Scott Librarian at the University of Alberta; TP, and a Registered Nurse with a master’s degree in library science employed at the University of Alberta was employed by our research team).

The search comprised three concepts based on the PICOS tool and then expanded upon using Medical Subject Headings (MeSH) and organized using appropriate Boolean search terms (Table 1). Concepts used in the search were related to caregivers, pediatric HF, and health information needs and experiences. To ensure thoroughness during the title-abstract screening phase, studies that focused on caregivers caring for a child with congenital heart disease were automatically carried into the second full-text screening phase, as children with congenital heart disease are all susceptible to experiencing HF. The full-text article would capture any reference to caregiver experiences in the chronic HF context nested within these studies. In anticipation of limited qualitative studies being retrieved, we applied no restrictions concerning time, country, or study design during the search.

**Table 1.**
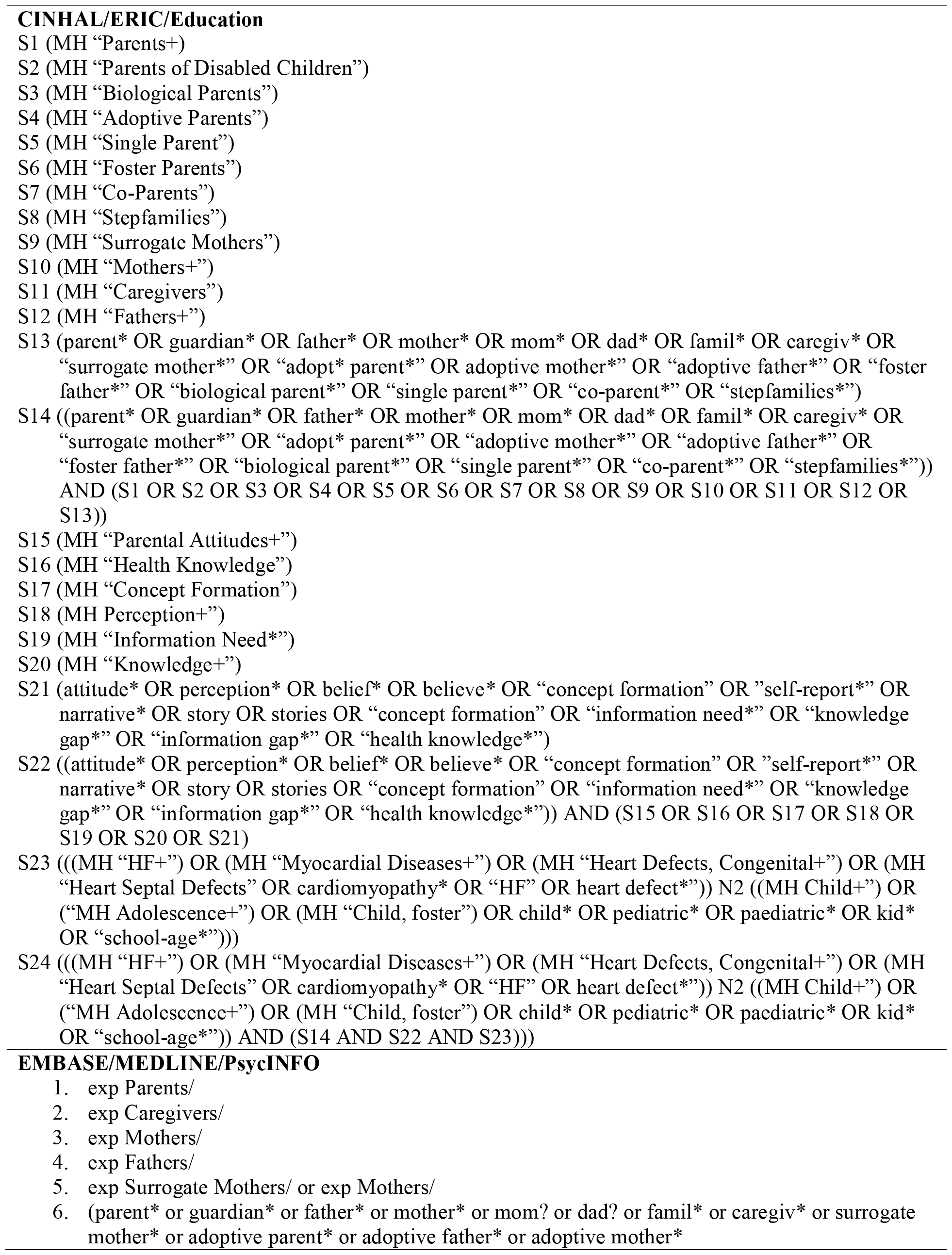
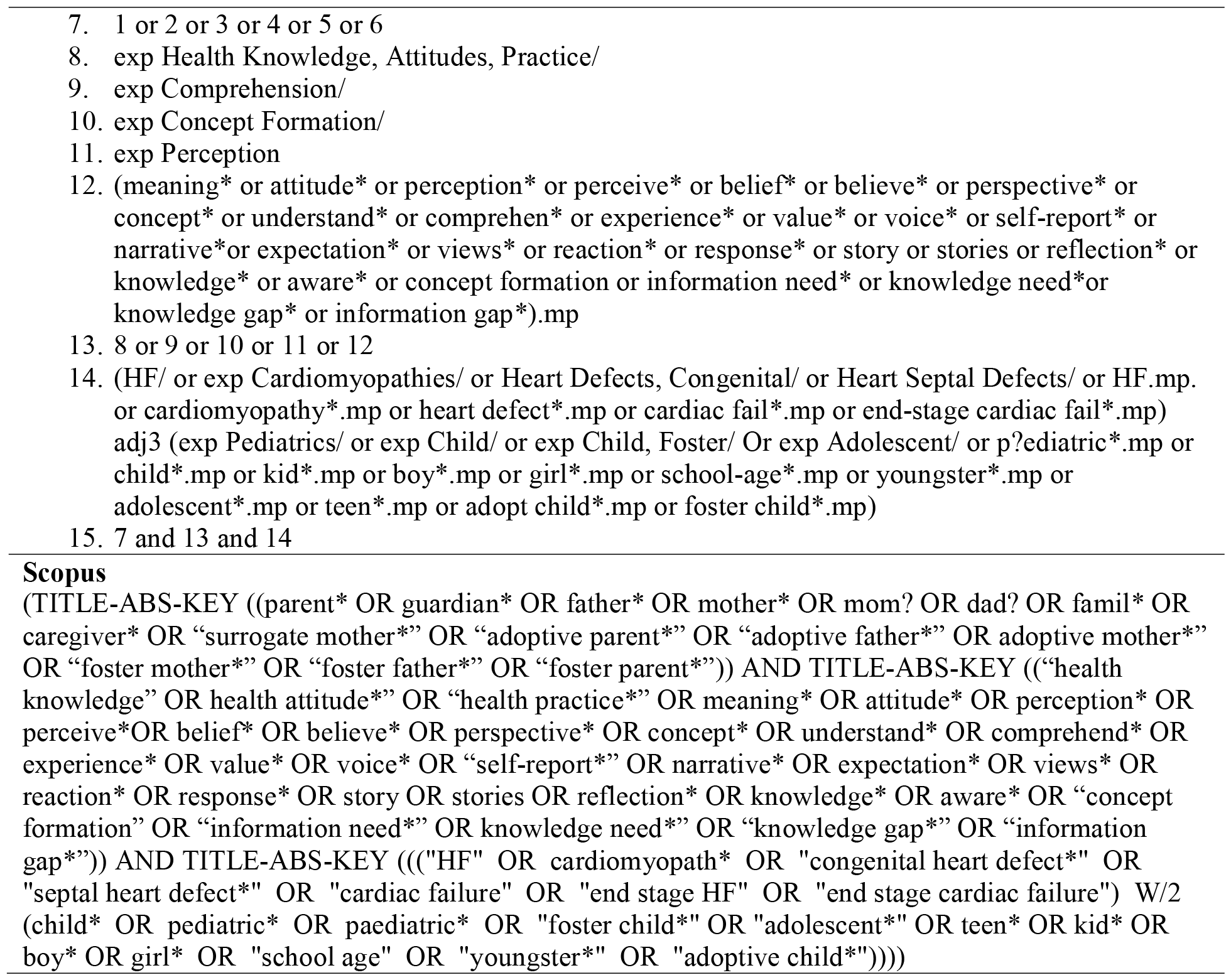
Search Strategies (organized by database)

### 2.4 Study selection

Search results were saved into the EndNote 21 reference manager and then imported into Microsoft Excel by the primary reviewer (CC). A senior researcher guided the entire synthesis process (SDS). The primary and secondary screeners (CC, KS) have several years of clinical nursing experience in chronic children’s HF, providing a strong foundation for decision-making during the inclusion/exclusion process.

The reviewer (CC) verified the imported list for accuracy by data validating all columns and confirming available and correct abstracts. To keep screening an independent process, The primary reviewer (CC) uploaded an Excel spreadsheet onto a Google Drive spreadsheet with separate tabs for the second reviewer (KS).

First, titles and abstracts were screened. The inclusion and exclusion criteria guide was built into the uploaded Excel spreadsheet by the primary reviewer (CC) using the data validation function to avoid any extraneous text answers. Studies that did not automatically import abstracts into the Excel spreadsheet were searched and included by the primary reviewer (CC) or automatically included in the full-text review list. If a study related to pediatric congenital heart disease or any other disease state that could result in a child having heart failure symptoms, it was automatically included in the final full-text screen for more in-depth consideration. All children with congenital heart disease are at risk of developing HF symptoms and themes could have been embedded into any study.

Both reviewers assessed all studies for inclusion and exclusion using the ‘Title/Abstract Inclusion/Exclusion Screening Guide & Definitions: PICOS Tool’ (Table 2) developed by the primary reviewer (CC). To ensure clarity and consistency regarding inclusion and exclusion, the primary reviewer (CC) compared the first 20 responses provided by both reviewers for accuracy before the rest of the screening took place. During this stage, no discrepancies with decisions needed to be resolved with the senior author (SDS). The reviewers discussed two studies at this stage (CC, KS) with resolution.

**Table 2:**
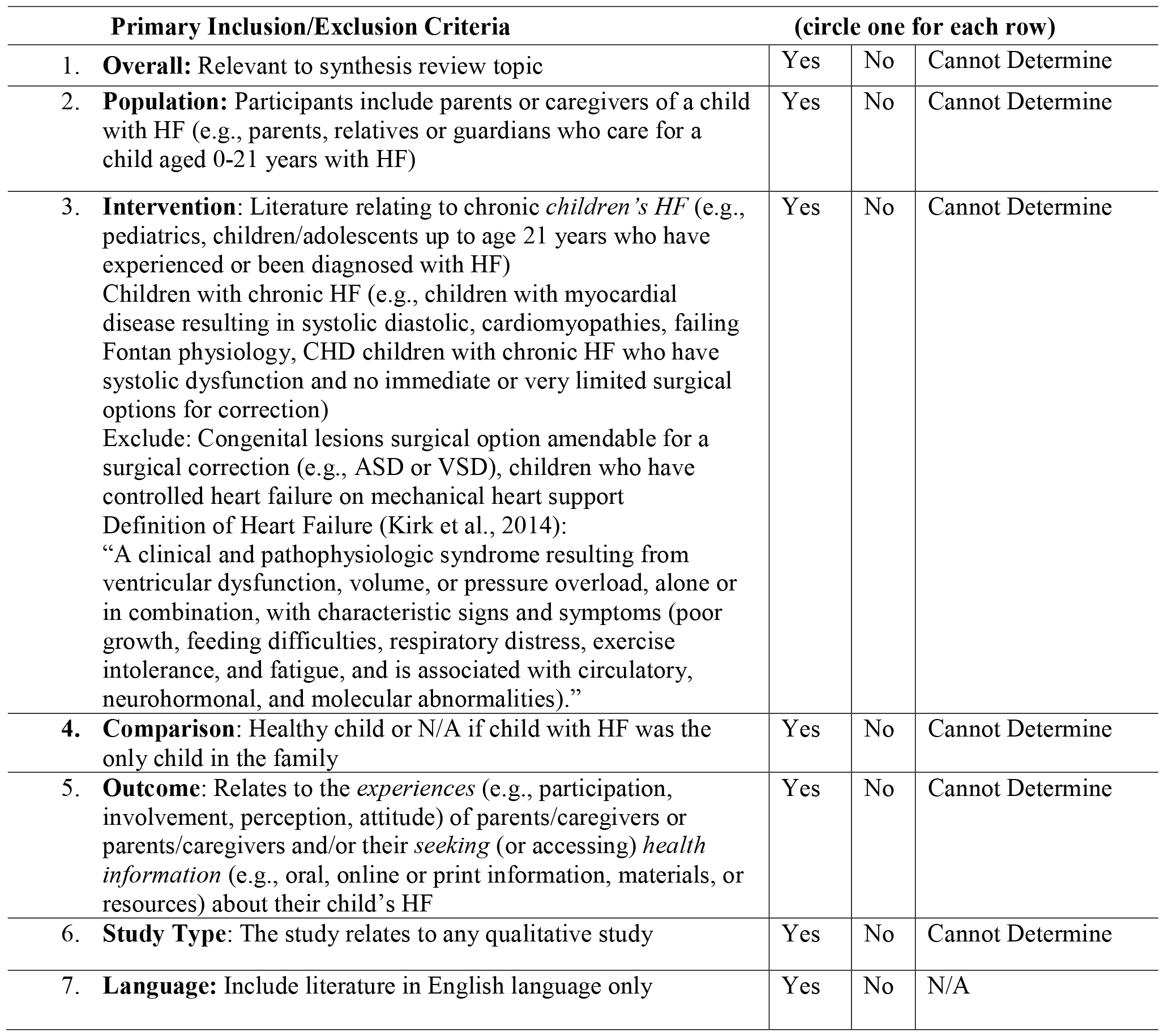
Inclusion/Exclusion Guide. **Inclusion/Exclusion Screening Guide & Definitions: PICOS Tool** **Title:** A Qualitative Evidence Synthesis Exploring Parent Information Needs and Experiences Caring for a Child with Chronic Heart Failure **Research Question:** What are the experiences and information needs of parents or caregivers who care for a child with heart failure?

### 2.5 Quality Assessment & Data Extraction

The Joanna Briggs Institute (JBI) Appraisal Checklist for Qualitative Research is a concise 10-question list that assesses for methodological quality using a structured four-point Likert scale format (e.g., yes, no, unclear, not applicable).^14^ The checklist has been used in prior studies, is available online and is a coherent tool with straightforward questions for reviewers.^15^ This stage was completed independently by each researcher (CC, KS) (Table 5). Critical appraisal results inform the synthesis stage by acknowledging strengths or biases in the currently published data, identifying key areas for growth and recommendations for future research. As Sandelowski and Barroso (2007) outlined, no studies would be excluded from our review due to poor quality due to the nature of qualitative research.^11^

### 2.6 Analysis

Our method of analysis was to complete Sandelowski and Barroso’s (2007) two-stage metasummary followed by their meta-synthesis. This method employs a two-stage approach to generate detailed effect sizes and novel interpretations of all previous published qualitative studies,^11^ while reducing the third-order analysis issues of stripping key contextual data.^15^ This analysis process results in a deeper, more coherent description of a specific qualitative phenomenon, uncovering new findings from primary studies.^11^

## 3.0 RESULTS

The PRISMA flow diagram outlines the inclusion-exclusion process (Figure 1). After de-duplication, 2,425 studies were identified. Thirty-nine studies made it to the full-text screening stage. One study met the inclusion criteria. A detailed summary of the rationale is included in Figure 1.

**Figure 1.**
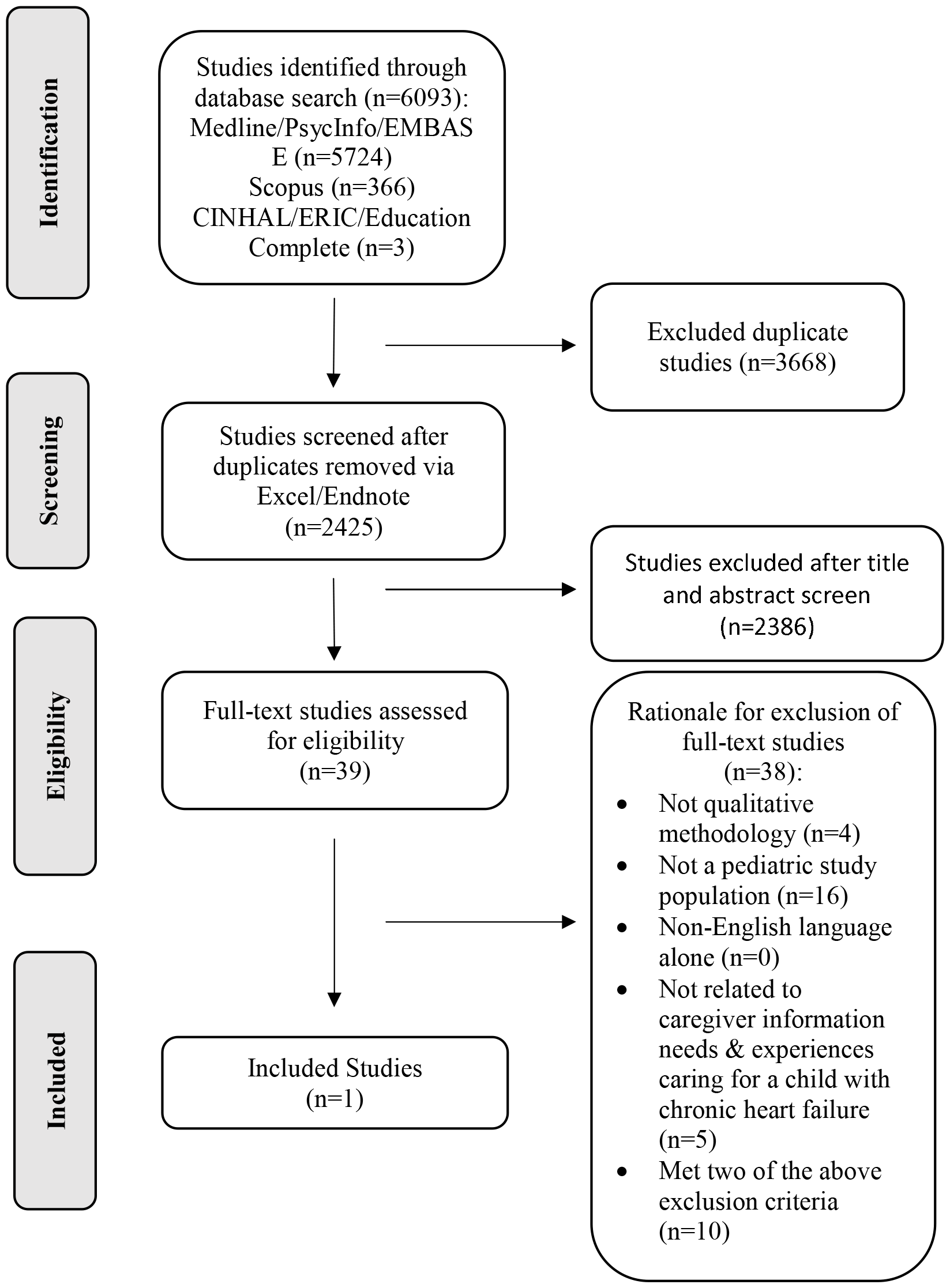
PRISMA Flow Diagram

The included study (Zhang et al., 2023) was conducted in China from April 2021 to 2022 and published in English. The study explored the experience of family management among caregivers who have a child with chronic HF, recruiting caregivers and interviewing them using semi-structured interviews.^16^ The qualitative study employed a descriptive phenomenology lens to keep findings as close to the data as possible. The authors stated this was a congruent method for the study design as they sought to understand the family’s experience of managing a child with heart failure as the environment and personal relationships impact it rather than interpret it. Data was analyzed using Colaizzi’s seven-step analysis. The findings included three themes and 10 sub-themes (Table 4).

**Table 3.** Study Characteristics.

| **Author/Year/Title/DOI** | **Study Design/Aim** | **Classification of Findings**<br>**(Defined by Sandelowski & Barroso, 2007)** | **Location/Context** | **No. of Participants/Reported Sexes/Race & Ethnicity** |
| --- | --- | --- | --- | --- |
| A. Z. Zhang, X., Shen, Q., Zhang, Q., Leng, H.<br>2023<br>Family management experience of parents of children with chronic heart failure: A qualitative study<br>doi.org/10.1016/j.pedn.2023.07.006 | Husserl's Phenomenological Theory (Descriptive Phenomenology) using semi-structured interviews (10 face-to-face, 6 WeChat video)<br>Duration of each interview: 20 - 60 minutes<br>Aim: This study explored the experience of family management from the perspective of the parents of children with CHF and may provide a reference for pediatric nursing staff to develop family management intervention programs. | Conceptual thematic description (bordering left on the grey towards thematic survey) | Parents of hospitalized children with CHF in the cardiovascular department in Chongqing, China from April 2021 to 2022 | 16 parents (purposive Sampling), no breakdown of male vs. female<br>Sex not reported, only identity (13 mothers, three fathers)<br>Race & Ethnicity: Not reported<br>Participant Ages (Years):<br>Overall Parent Age Range: 25 - 56<br>Mothers Age Range: 25 - 48 (mean 36.8)<br>Father Age Range: 33 - 56 (Mean 42) |

**Table 4.** Study Reported Outcomes.

| Theme #1: Weakened Family Socialization | Theme #2: Experience of Five Psychological Stages | Theme #3: Family Management Dilemmas |
| --- | --- | --- |
| 2 Sub-themes:<br>**1. Diminished Parental Role in Social Education:** Some parents react with a mindset of compensating for the child, resulting in a lack of restraint regarding the child's behaviour and guidance related to the child's emotional regulation. Over time, children with CHF experience a range of psychological, behavioural, and social adaptation problems.<br>**2. Insufficient Socialization of Children:** Children with CHF have delayed growth and development and decreased activity endurance, which limits or even blocks their social activities. | 5 Sub-themes:<br>**1. Exhaustion:** In the process of medical treatment, family management, and uncertainty, parents often experience psychological pressure and even criticism, thus making them sensitive to sadness and Resistance: Little knowledge about the disease among parents made it difficult for parents to accept the reality of their children's illness at the time of initial diagnosis.<br>**2. Self-blame:** When parents begin to acknowledge that their children are ill, some of them feel guilty and blame themselves for their children's illnesses. Similarly, during medical treatment, unintentional blame by people nearby can intensify this sense of self-blame.<br>**3. Worry:** Parents' desire for medical treatment increased after they adjusted their emotions. During this time, parents were worried about their child's prognosis and future.<br>**4. Exhaustion:** In the process of medical treatment, family management, and uncertainty, parents often experience psychological pressure and even criticism, thus making them sensitive to sadness and even exhaustion.<br>**5. Acceptance:** During a protracted time of care, parents started to accept the significant changes the disease had caused in the family. Several parents lost confidence and felt helpless. Nonetheless, other parents persisted in changing their perspective, regaining their confidence, and embracing their children's illnesses with optimism. | 3 Sub-themes:<br>**1. Low Social Awareness of the Disease:** Awareness of CHF among parents, the public, and grassroots pediatric nursing staff is low, and that creates many challenges in the family management of the disease. Parents have inadequate disease management ability, which is mainly manifested in insufficient medication management at home, irregular management, such as a low-salt diet, and a lack of scientific disease monitoring techniques.<br>**2. Heavy Economic Burden:** When a child becomes unwell, the family's financial burden increases due to the cost of medical care. One parent must quit his/her job to care for the sick child at home, which reduces economic resources. The disease treatment is a long process, and the expenditure is unpredictable, which makes the economic situation in the family extremely burdensome.<br>**3. Limited Coping Styles:** During the entire process, the parents tried to keep the family stable but often ignored their health and other children's feelings. |

**Table 5.**
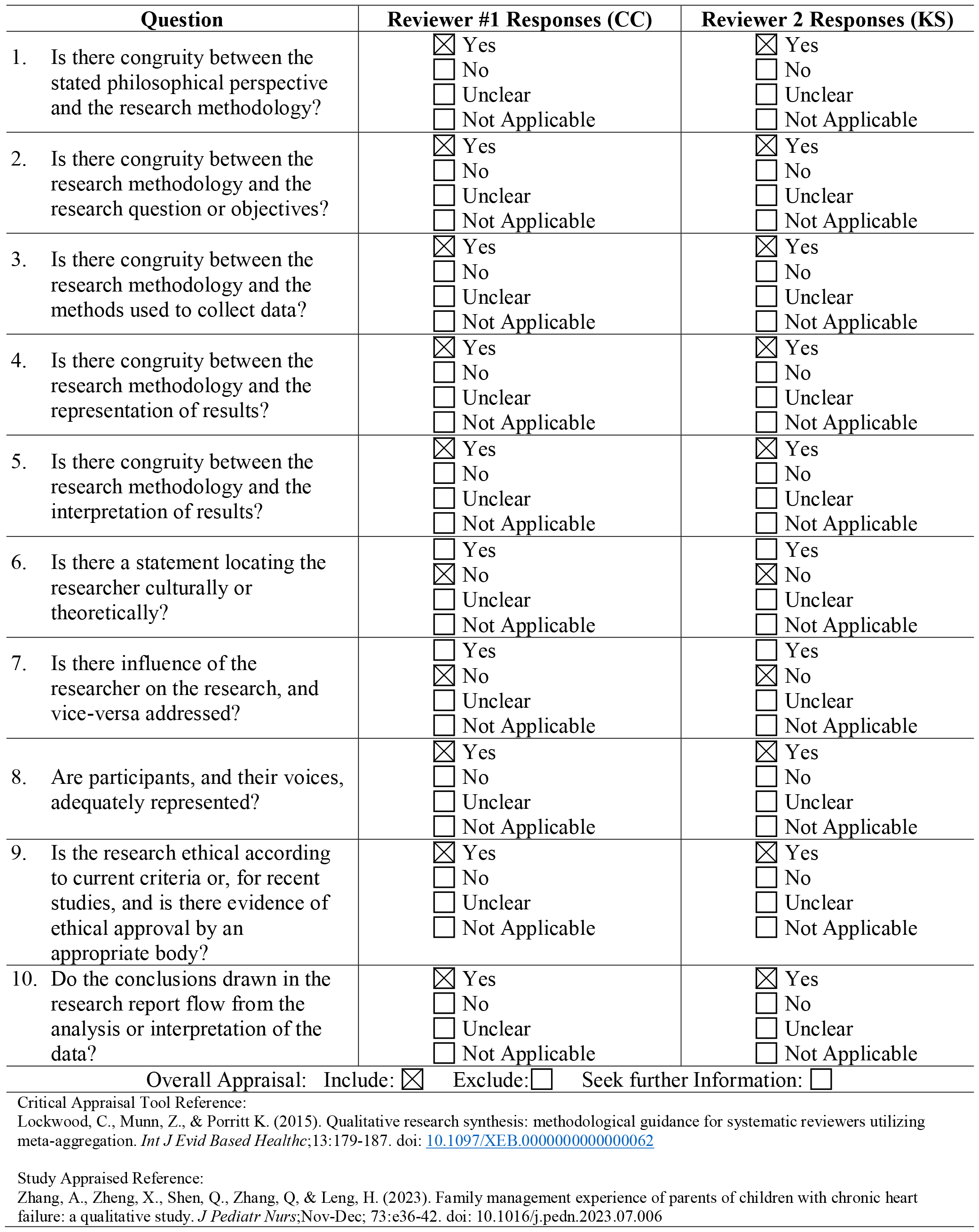
JBI Quality Appraisal Checklist.

| Question | Reviewer #1 Responses (CC) | Reviewer 2 Responses (KS) |
| --- | --- | --- |
| 1. Is there congruity between the stated philosophical perspective and the research methodology? | ☒ Yes<br>☐ No<br>☐ Unclear<br>☐ Not Applicable | ☒ Yes<br>☐ No<br>☐ Unclear<br>☐ Not Applicable |
| 2. Is there congruity between the research methodology and the research question or objectives? | ☒ Yes<br>☐ No<br>☐ Unclear<br>☐ Not Applicable | ☒ Yes<br>☐ No<br>☐ Unclear<br>☐ Not Applicable |
| 3. Is there congruity between the research methodology and the methods used to collect data? | ☒ Yes<br>☐ No<br>☐ Unclear<br>☐ Not Applicable | ☒ Yes<br>☐ No<br>☐ Unclear<br>☐ Not Applicable |
| 4. Is there congruity between the research methodology and the representation of results? | ☒ Yes<br>☐ No<br>☐ Unclear<br>☐ Not Applicable | ☒ Yes<br>☐ No<br>☐ Unclear<br>☐ Not Applicable |
| 5. Is there congruity between the research methodology and the interpretation of results? | ☒ Yes<br>☐ No<br>☐ Unclear<br>☐ Not Applicable | ☒ Yes<br>☐ No<br>☐ Unclear<br>☐ Not Applicable |
| 6. Is there a statement locating the researcher culturally or theoretically? | ☐ Yes<br>☒ No<br>☐ Unclear<br>☐ Not Applicable | ☐ Yes<br>☒ No<br>☐ Unclear<br>☐ Not Applicable |
| 7. Is there influence of the researcher on the research, and vice-versa addressed? | ☐ Yes<br>☒ No<br>☐ Unclear<br>☐ Not Applicable | ☐ Yes<br>☒ No<br>☐ Unclear<br>☐ Not Applicable |
| 8. Are participants, and their voices, adequately represented? | ☒ Yes<br>☐ No<br>☐ Unclear<br>☐ Not Applicable | ☒ Yes<br>☐ No<br>☐ Unclear<br>☐ Not Applicable |
| 9. Is the research ethical according to current criteria or, for recent studies, and is there evidence of ethical approval by an appropriate body? | ☒ Yes<br>☐ No<br>☐ Unclear<br>☐ Not Applicable | ☒ Yes<br>☐ No<br>☐ Unclear<br>☐ Not Applicable |
| 10. Do the conclusions drawn in the research report flow from the analysis or interpretation of the data? | ☒ Yes<br>☐ No<br>☐ Unclear<br>☐ Not Applicable | ☒ Yes<br>☐ No<br>☐ Unclear<br>☐ Not Applicable |
| Overall Appraisal: Include: ☒ Exclude: ☐ Seek further Information: ☐ |  |  |

Critical Appraisal Tool Reference:
Lockwood, C., Munn, Z., & Porritt K. (2015). Qualitative research synthesis: methodological guidance for systematic reviewers utilizing meta-aggregation. *Int J Evid Based Healthc*;13:179-187. doi: [10.1097/XEB.0000000000000062](https://doi.org/10.1097/XEB.0000000000000062)
Study Appraised Reference:
Zhang, A., Zheng, X., Shen, Q., Zhang, Q., & Leng, H. (2023). Family management experience of parents of children with chronic heart failure: a qualitative study. *J Pediatr Nurs*;Nov-Dec; 73:e36-42. doi: 10.1016/j.pedn.2023.07.006

### 3.2 Classifying the Findings

Sandelowski and Barroso (2007) situate the findings of included studies on a continuum to understand how researchers analyzed the primary data, entitled classifying the findings. The continuum indicates the degree of data transformation during the analysis phase (e.g., level of interpretation). The continuum runs from left to right, with the left side being the closest findings to the participant’s descriptions (topical/survey data) to the far right (conceptual description/interpretative findings). This process forces the reviewers to truly evaluate the findings through a more critical lens, considering and selecting a description in Sandelowski and Barroso’s constructed scale rather than just merely restating the methods claimed by the authors.

This classification of this study was conceptual/thematic. Their study aimed to stay close to the data to highlight caregivers’ experiences with a marginal amount of interpretation from the researchers using a qualitative descriptive phenomenology approach. The authors included only one or two minimally interpretive sentences in each subtheme and let numerous rich quotes for each subtheme speak for themselves.

### 3.3 Quality Assessment & Data Extraction

Study characteristics are included in Table 3, highlighting key details. Sixteen participants (13 mothers and three fathers) were recruited. Data was collected through in-person and online semi-structured interviews ranging from 20-60 minutes (10 in-person and six recorded WeChat interviews). Transcripts were transcribed verbatim by the study team shortly after each interview and analyzed using content analysis. Three themes with 1 to 5 subthemes nested within each category. Theme titles were 1) weakened family socialization, 2) experience of five psychological stages, and 3) family management dilemmas (Table 4).

Both reviewers (CC, KS) completed the quality appraisal using the Joanna Briggs Institute (JBI) qualitative checklist.^14^ The concise 10-item checklist evaluates a study’s rigour, epistemology, and ontology.^18^ Sandelowski and Barroso (2007) suggest that the purpose of critical appraisal is to provide information about the quality of the evidence rather than exclude the papers, as exclusion may introduce bias.^11^ Furthermore, widespread debate exists about rigour within qualitative research due to a lack of consensus, so it is more cautious about including all qualitative studies.^11^

Both reviewers scored the study and unanimously agreed this study had many rigorous qualities (Table 5). The study provided excellent congruity between the philosophical perspective, research methodology, and objectives. Ethics were also considered and obtained. The only aspects not stated in the study were elements relating to locating the researcher culturally or theoretically and their influence on the researcher’s findings, which may have introduced some bias. Overall, this study withheld a high level of methodological quality.

### 3.4 Metasummary and Metasynthesis

No synthesis could be completed since only one study was included in our review. It was impossible to complete either the metasummary (e.g., effect size calculation) or metasynthesis (e.g., new interpretations of the included studies) steps outlined in Sandelowski and Barroso’s handbook.

## 4.0 DISCUSSION

The primary objective of this qualitative synthesis was to uncover and synthesize literature relating to caregivers’ information needs and experiences for a child with chronic HF. To our knowledge, no previous synthesis has been completed on this topic. Our research uncovered only one recent qualitative study by Zhang et al (2023) that met our inclusion criteria, confirming a significant knowledge gap about caregivers’ information needs and experiences caring for a child with HF. The study included 16 participants, uncovering three themes and ten subthemes relating to caregivers’ family management who have a child with chronic heart failure (Table 3). The location of the study was in China, signifying that no North American literature was uncovered in our study, identifying a gap in North American knowledge gap.

The results from the included Zhang et al. (2023) study are not unexpected. The reviewers suspected it that families who have a child with chronic heart failure experience difficulties with socialization and economic burden, imposing psychological repercussions on caregivers. Caring for a child with such a complex condition is taxing on families and requires constant monitoring for adverse symptoms or clinical deterioration. Living in a state of constant uncertainty has negative effects on families. This paper also highlighted that participants mentioned a low social awareness of this disease due to its rare; caregivers who have children who are newly diagnosed experience feelings relating to a lack of educational information.

### 4.1 Nature of Chronic Heart Failure on Family Life

Caregivers are responsible for essential daily management tasks for managing their children with HF. Healthcare providers spend countless hours training to learn how to care for children with HF; therefore, a parallel focus on caregivers’ education and training in this setting is not and should be prioritized for parents to learn such complex knowledge. But despite this, we uncovered a significant knowledge gap about parents’ information needs and experiences relating to their child’s heart failure from this one study, which is troubling given the pressures placed on parents. Caregivers are expected to understand and become proficient in highly complex medical knowledge about their child’s HF and make day-to-day decisions about multiple medications, fluid restrictions, early symptom recognition, complex cardiology terminology, and specialized medical therapy regimes (e.g., high-calorie and low-sodium diets, feeding via nasogastric and g-tubes, central line care and juggling multiple healthcare provider appointments).^4^ Caring for a child with chronic HF differs from caring for a child with CHD. Children and families have the opportunity for surgical correction or palliation, bringing rise to unique experiences that are different from children who experience chronic heart failure symptoms. Examples include surgical complications or experiences related directly to surgical experiences (e.g., cancellations, surgical preparation, meeting new team members, and different information needs based on their cardiac lesion).^18,19^

### 4.2 Caregiver Experience in Adult HF

Since our review focused on caregiver information needs and experiences, literature on adult HF caregiver experience was excluded but better studied. Given children’s lack of critical thinking skills and reliance on caregivers to provide care due to their immature growth and development, there is limited opportunity to generalize and extrapolate adult HF literature to the pediatric population. Not surprisingly, a state of science review by the American Heart Association suggests that unpaid support from family and friends of adults with HF imposes a high level of strain on their caregivers.^20^ They also report that caregivers can experience feelings of doubt and anxiety, the need for constant guidance and support from healthcare providers, unmet personal needs, the continuous juggling of caregiving tasks, and the continual adaptation of strategies to normalize their lives, similar to the findings from Zhang’s study.

Kitko et al. (2020) also propose that advancements in adult HF treatments in the home setting have become more intensive and increasingly precarious for the adult HF patient population to be delivered by family members. This is likely true in the pediatric population as they have a healthy adult to care for them. Tasks now performed in the home are ones that healthcare professionals in clinical settings would typically provide in the past, signifying that patients are being discharged into the community earlier. Adult HF caregivers spend an average of 22 hours per week,^20^ which we know in pediatrics will be higher given children’s sole reliance on their caregivers for all or most of their day-to-day care and decision-making. Caregiver needs in the adult context have been extensively documented compared to pediatrics, identifying support relating to daily living, improving, and maintaining self-care, psychological support, and navigating the complex medical system. Kitko et al. (2020) also highlight that caregivers handle some of these tasks simultaneously, requiring an increased ability to think critically. They also suggest that caregivers’ roles are invaluable in preventing costly hospital readmissions, which will unarguably be a factor in pediatrics.

Lastly, a qualitative study by Sedlar (2020) completed qualitative interviews with adult HF patients, their informal caregivers, and healthcare providers.^21^ Informal caregivers felt they mostly took on practical tasks (e.g., medication administration and meal preparation), and 33% of caregivers reported providing emotional support to their spouses with HF.^21^ Half of the informal caregivers reported experiencing anxiety related to the future and their ability to manage sudden deterioration. Most (90%) of informal caregivers reported changes in their family roles and relationships after the diagnosis, forcing them to change their lifestyle to adapt to the patient’s limitations. Notably, a third of the caregivers described their needs as less important than their spouse’s needs with HF. Thankfully, two-thirds of informal caregivers felt acknowledged and ‘part of the team’ at a medical appointment, which we hope to find in our next research study involving qualitative pediatric interviews.

It is anticipated that many of these themes within the adult literature would have been uncovered if qualitative studies in the pediatric context were included. However, there was little knowledge in the single-included study about caregivers’ information needs that we need to build on in future research. Like Kitko (2020), the included study reported similar findings of caregiver anxiety, self-doubt, financial strain, and unmet personal needs, like the pediatric lens presented in Zhang’s study.

### 4.3 The Importance of Knowledge Gaps

Since our study identified a single study with 16 participants, only introductory knowledge exists on this complex topic. While our findings share limited findings, this is similar to key characteristics of reviews that uncover no studies, called empty reviews.^22,23^ Empty reviews were first mentioned by Lang et al. (2007), who suggested they are rare.^23^ Yaffe et al. (2012) conducted a systematic review in the Cochrane Database for Systematic Reviews pertaining to the frequency of empty reviews, concluding that they were not as rare as once thought as they occurred in 1 in 10 reviews.^22^ Arguments in the literature state that empty reviews may appear to offer no conclusions as there is no evidence, leading to an overall general disappointment in the absence of recommendations or guidance.^22,23^ However, alternative arguments support the importance of empty reviews because authors acknowledge a critical knowledge gap exists for a specific topic, validating the need to prioritize future research.^24^ So, while no synthesis could be conducted, this finding highlights the need for further research on this important topic.

### 4.4 Limitations of this Study

Like all studies, this study is not free of limitations. Given the nature of the design, only qualitative manuscripts were included. This approach aligned with the qualitative nature of the research question relating to caregivers’ information needs and experiences, which cannot be appropriately addressed using only quantitative methods. Therefore, the decision to only include qualitative papers was made to uncover and synthesize detailed, contextual descriptions of caregivers’ and caregivers’ relating to their information needs and experiences.

The second limitation pertains to the reviewers’ (CC, KS) anticipation and perhaps confirmation bias. The reviewers with advanced clinical knowledge through years of practice experience in pediatric chronic HF anticipated there would be limited to no included studies. To ensure confidence in our search strategy, no study type limitation was applied in the initial search, in addition to having the privilege of consulting two librarians. Studies were excluded independently and discussed after each screening phase to ensure no knowledge of our topic was missed. We also hand-searched the included studies’ reference list to extend our search for included studies.

## 5.0 CONCLUSION

We uncovered two key findings through this qualitative evidence synthesis. First, this review was the first of its kind in the rapidly evolving field of pediatric HF, looking to understand caregivers’ information needs and experiences caring for a child with HF. Secondly, only one study met our inclusion criteria, highlighting a considerable knowledge gap in the literature that needs further exploration. It is known that guidelines have been developed to advance treatments within the last decade; however, our review demonstrates that research exploring caregivers’ information needs and experiences has been given little attention despite providing life-saving treatments to these vulnerable children.

## Data Availability

The data that support the findings of this study are available from the corresponding author, Shannon D. Scott, upon reasonable request.

## Statements

## Acknowledgements/credits/disclaimers

Thank you to Megan Kennedy, MLIS from the John Scott Library at the University of Alberta, for her assistance in formulating and developing our initial robust search strategy for this review.

